# A Digital Biomarker Dataset in Hematopoietic Cell Transplantation: A Longitudinal Study of Caregiver-Patient Dyads (dHCT)

**DOI:** 10.1101/2024.11.21.24317641

**Authors:** Aditya Jalin, Nawat Swatthong, Michelle Rozwadowski, Rajnish Kumar, Deb Barton, Tom Braun, Noelle Carlozzi, David A. Hanauer, Afton Hassett, Sung Won Choi

## Abstract

**Background:** Hematopoietic stem cell transplantation (HCT) is a potentially life-saving therapy for individuals with blood diseases, but involves a challenging recovery process that requires dedicated caregivers. The complex interplay between emotional distress, care partner (or unpaid caregiver) burden, and treatment outcomes necessitates comprehensive physiological and psychological measurements to fully understand these dynamics.

**Findings:** We collected longitudinal data from 166 HCT caregiver-patient dyads over 120 days post-transplant as part of a randomized controlled trial (NCT04094844). Data were gathered using the Fitbit® Charge 3 device, a custom mood-reporting app with positive psychology-based activities (Roadmap), PROMIS® health measures, and clinical events. The dataset includes minute-level heart rate, daily sleep metrics, step counts, self-reported mood scores, app usage metrics, PROMIS® T-scores (i.e. global health, depression), infection and readmission records, and clinical outcomes (e.g., acute and chronic graft-versus-host disease, relapse, mortality). Physiological data were available for both caregivers and patients. Data validation confirmed high compliance with mood reporting and the presence of physiological patterns between caregivers and patients that differed (i.e., lower activity in patients compared with caregivers across time).

**Conclusions:** This dataset offered an unprecedented view into the daily fluctuations of caregiver and patient well-being throughout the critical post-HCT period. It provided a valuable resource for researchers investigating the impact of mHealth app interventions, including emotional distress and physiological markers on treatment course and clinical outcomes. Our unique dataset informs interventions that may address caregiver support, patient care, or dyadic-focused strategies and enable novel analyses of single member or dyadic dynamics in HCT treatment.

## Context

Hematopoietic stem cell transplantation (HCT) is a complex and potentially life-saving procedure used to treat a variety of hematologic malignancies, genetic disorders, and immune system deficiencies[1]. The procedure involves infusing healthy stem cells to replace damaged or diseased cells in the patient’s bone marrow. While HCT can significantly improve long-term survival rates, it is an intensive treatment that places enormous physical, mental, and emotional toll (i.e., health-related quality of life [HRQOL]) on both patients and their caregivers (*dyads*) [2,3].

The post-transplant period is crucial for patient recovery due to the constant (24/7) care and monitoring of potential life-threatening complications, such as graft-versus-host disease (GVHD), infections, and relapse [4,5]. This intense caregiving process can lead to substantial burden and stress for family caregivers, which may adversely impact their own health and well-being, as well as the outcomes of the patients they care for [3,6,7].

The complex interplay between caregiver well-being and patient outcomes in HCT has been recognized as an important area of study [8,9]. However, most existing research relies on isolated snapshots of caregiver and patient status, often using retrospective self-reports or infrequent assessments. These methods fail to capture the dynamic nature of the caregiving experience and may miss important fluctuations in physiological and psychological states [10].

Furthermore, there is growing interest in leveraging mobile health (mHealth) technologies to support caregivers and patients through this challenging process [11,12]. Wearable devices and smartphone apps offer the potential for continuous, real-time monitoring of physical, mental, and emotional well-being, providing a more comprehensive view of the HCT experience.

In response to these gaps in knowledge and emerging technological opportunities, we designed this study to collect a comprehensive, longitudinal dataset that captured both physiological and psychological measures from HCT caregiver-patient dyads over a 120-day post-transplant period. By incorporating continuous monitoring through wearable devices (Fitbit® Charge 3[13]), a custom mHealth app (Roadmap[14]), standardized health measures (PROMIS®[15]),and clinical events (i.e., infection and readmission records, acute and chronic graft-versus-host disease, relapse, mortality), this dataset provides an unprecedented view into the daily fluctuations and interdependencies of caregiver and patient well-being throughout the critical post-HCT period.

The overarching purpose of generating this rich dataset was multifold:

1. To better understand the dynamics of caregiver burden in HSCT, including how burden changes over time and how it relates to patient outcomes.
2. To explore the potential benefits of positive psychology interventions delivered via mobile technology.
3. To investigate the relationships between physiological markers, psychological well-being, and clinical outcomes in both caregivers and patients.
4. To provide a comprehensive resource for developing machine learning models that could predict adverse events or identify periods of high stress in real-time.

This dataset builds upon previous studies in caregiver research and mHealth interventions, offering a unique resource for researchers to delve deeper into the complex realities of the HCT experience. It has the potential to inform the development of more targeted, effective support strategies for both caregivers and patients, ultimately improving the HCT journey for all involved [6].

By making this dataset available to the research community, we specifically aim to (i) accelerate progress in understanding and supporting the HCT process; (ii) promote the development of innovative digital health interventions; and (iii) contribute to the growing field of dyadic health research in oncology and beyond.

## Methods

### Study Design

The full IRB-approved and registered protocol for this study was detailed in a previous publication[16]. What follows here is a summary of the key aspects of the study design relevant to understanding and navigating the rich dataset.

This study employed a randomized controlled trial (RCT) design to evaluate the effectiveness of a mobile health intervention (Roadmap) on caregiver HRQOL in the context of HCT. The study was conducted at the University of Michigan (U-M) Blood and Marrow Transplant (BMT) Program and recruited 166 caregiver-patient dyads undergoing allogeneic HCT, a sample size determined through power analysis to detect a clinically meaningful difference in the day 120 post-transplant PROMIS® Global Health score in caregivers [17].

Eligible caregivers were identified by patients as their primary caregiver, providing at least 50% of care needs, a threshold consistent with previous caregiver studies[18]. Both caregivers and patients were required to be at least 18 years old, able to read and speak English, and capable of providing informed consent, consistent with standard ethical guidelines for clinical trials [19]. Patients were eligible if they were scheduled to undergo HCT, able to identify an eligible caregiver, and capable of signing the University of Michigan Caregiver Responsibility Contract along with their caregiver. Exclusion criteria were limited to patients not meeting the eligibility criteria for HCT at the U-M BMT Program, consistent with established U-M clinical practice guidelines (CPG) [20].

Figure 1 illustrates the comprehensive timeline of our study, from participant enrollment through data collection. Following consent, all enrolled dyads received Fitbit® Charge 3 devices and the Roadmap app, with data collection commencing on the day of transplant and continuing for 120 days. This included continuous physiological monitoring via Fitbit® devices, daily mood assessments, and periodic PROMIS® surveys, while clinical outcomes were tracked for a full year post-transplant

**Figure 1:**
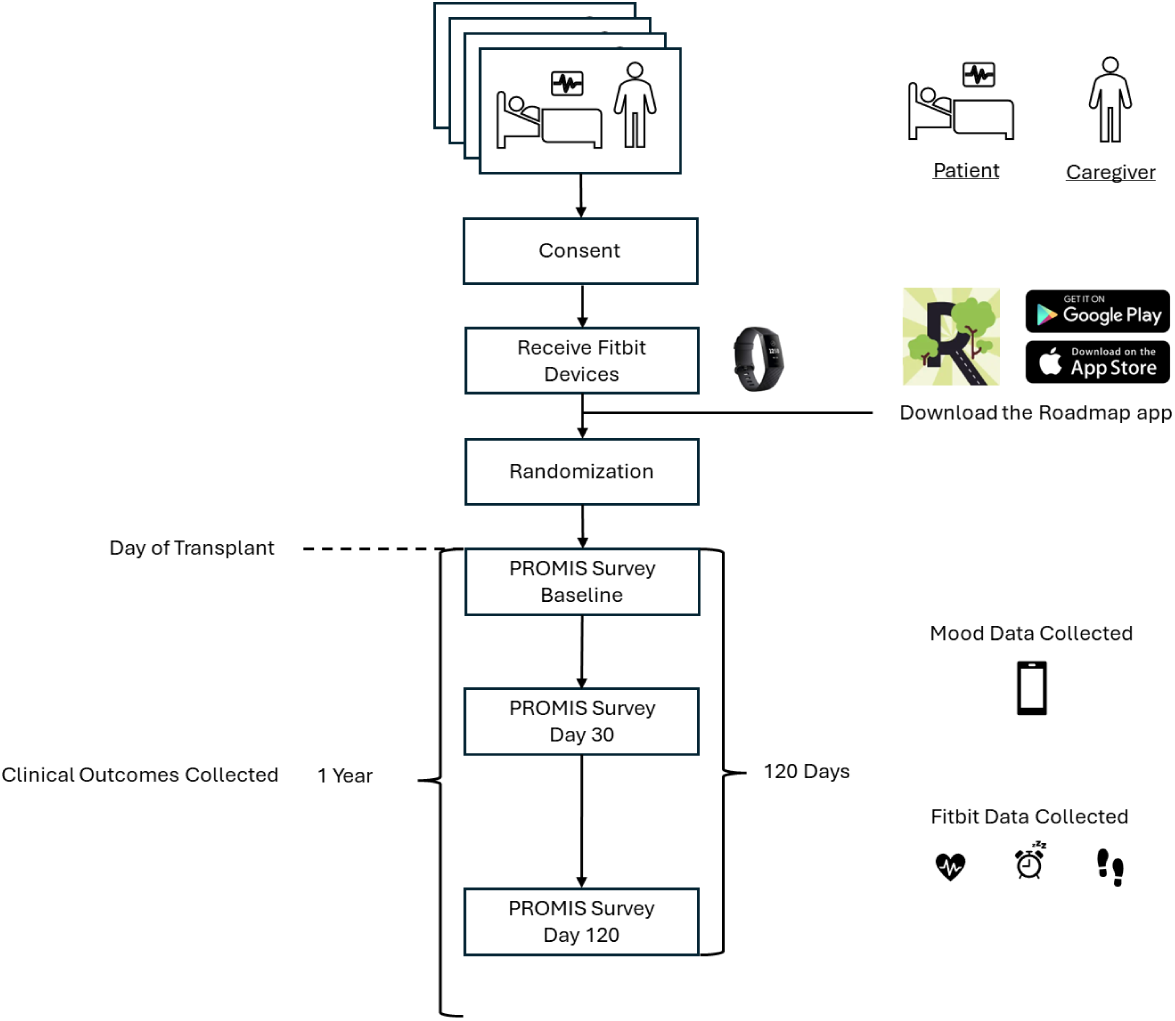
Study timeline and data collection framework of the study. Schematic representation of the study timeline and data collection process. Following consent, caregiver-patient dyads received Fitbit® Charge 3 devices and downloaded the Roadmap app. After randomization, data collection after the day of transplant, including PROMIS® surveys (baseline, day 30, and day 120), continuous Fitbit physiological measurements (heart rate, sleep, steps), and self reported daily mood. While the intensive monitoring period concluded at day 120, clinical outcomes continued to be tracked for one year post-transplant

Dyads were randomly assigned to one of two arms: the treatment arm, which received the Roadmap app with multicomponent features plus a Fitbit® Charge 3, or the control arm, which received the Roadmap app displaying physiological data only plus a Fitbit® Charge 3. As shown in Figure 2, all the patients only had access to mood reporting and Fitbit data visualization features, irrespective of intervention assignment. Only the caregivers in the intervention arm received access to positive psychology activities. A blocked randomization approach was used to ensure equal distribution between the two arms, overseen by the study statistician (TB). Caregivers were randomized no more than 21 days from the planned initiation of conditioning for transplant, and all patient treatments related to the transplant were scheduled prior to randomization to avoid potential biases.

**Figure 2:**
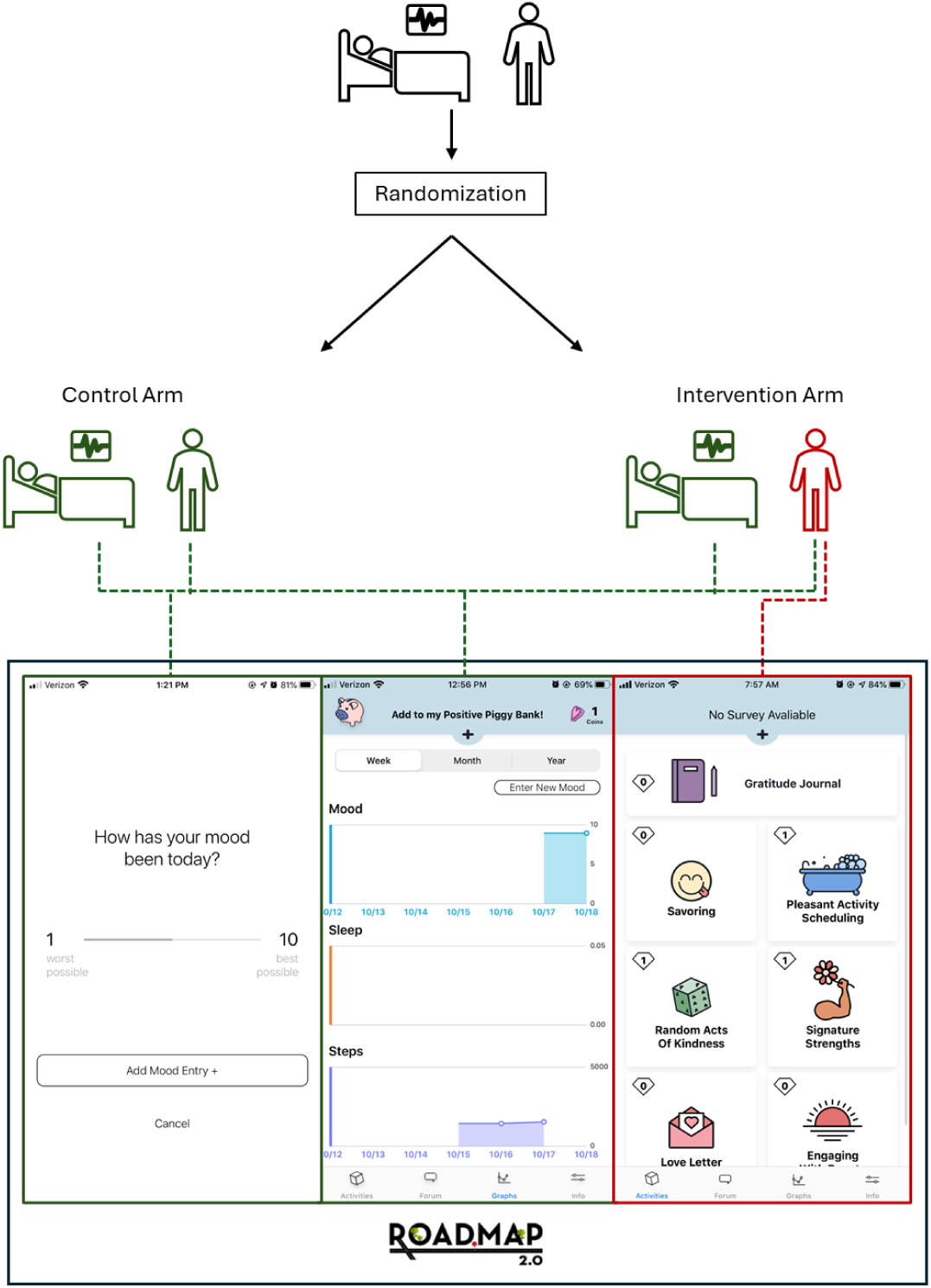
Overview of the study randomization and app functionality. Caregiver-patient dyads were randomized into control or intervention arms. The Roadmap app provided mood reporting and Fitbit data visualization features to all participants (shown in left and center screenshots). The positive psychology activities (3rd screen from the left) were exclusively available to caregivers in the intervention arm. Add activities in deep blue

This sampling and randomization strategy was designed to provide a comprehensive view of the HCT experience for both caregivers and patients. It allowed for analysis of the effectiveness of the Roadmap intervention on caregiver HRQOL, while controlling for potential confounding factors through randomization.

The longitudinal nature of the study, with data collection spanning 120 days post-transplant, enabled researchers to monitor for changes in physiological and psychological measures over time, providing insights into the dynamic biobehavioral nature of the HCT experience for both caregivers and patients. This approach aligned with calls in the literature for more comprehensive, long-term assessments of HCT outcomes [10,21].

### Data Collection

Our data collection process was designed to capture a comprehensive, multi-faceted view of the HCT experience for both caregivers and patients. We employed a variety of tools and techniques to gather data over a 120-day period following the transplant, enabling us to monitor for changes and patterns over time. The data were collected using a combination of the Roadmap app, Fitbit® Charge 3 devices, and self-report surveys. The following subsections outline the different modalities of data that were collected.

#### Wearable Device Data

Fitbit® Charge 3 devices were used to collect continuous physiological data from both caregivers and patients. The data were transferred from the smartwatch to the Fitbit® app installed on the patient’s phone via Bluetooth Low Energy (BLE). The app then transmitted data to Fitbit’s® information management system securely and without identifiable information. The device was worn on either wrist to give accurate measurements. The device was capable of measuring heart rate, exercise amount, and sleep levels. The Fitbit® API enabled sleep (total minutes/day) and physical activity (total step count/day) data to be visualized on the Roadmap app.

#### Heart rate

Heart rate data were collected with a sampling rate that ranged from 5 seconds to 1 minute. This variability in sampling rate was due to changes in the Fitbit® API during the study period. To maintain consistency in the data, the heart rate sampling rate was standardized to 1 minute by choosing the median values for the heart rate across each person-minute. This approach allowed us to capture the overall trend of heart rate changes while minimizing the impact of short-term fluctuations or potential measurement errors.

#### Sleep patterns

Fitbit® collects two types of sleep data. The first, referred to as “classic” sleep, primarily relies on movement detection. This approach assumes a period of sleep after detecting approximately an hour of stillness in the absence of heart rate data[22]. Furthermore, manual sleep entries, sleep recorder for less than 3 hours, and critically low battery results in “classic” sleep entries. However, it is worth noting that this is an outdated version of sleep measurement that Fitbit® is planning to phase out.

The newer version, “sleep stages” tracking, offers a more nuanced analysis. This advanced approach combines movement data with heart rate patterns to estimate different phases of sleep. By incorporating physiological data beyond just movement, this method provides a more accurate differentiation between sleep and wake states, as well as insights into the quality and stages of sleep (e.g., light, deep, and REM). The dual approach to sleep tracking enables us to analyze not just the quantity but also the quality of rest, which is crucial given the potential impact of the HCT process on sleep patterns for both caregivers and patients.

#### Physical Activity

Physical activity was recorded through two measurements provided by the Fitbit® API. First, it recorded the number of steps taken per minute, offering a precise measure of ambulatory activity throughout the day. These data can be used to assess overall activity levels, detect periods of increased movement, and identify patterns in daily routines.

Additionally, the level of activity was recorded ordinally, categorized as “sedentary,” “lightly active,” “moderately active,” and “very active.” This classification provided a contextualized understanding of the intensity of activities beyond just step count. For instance, it may distinguish between periods of low-intensity movement (like gentle walking) and high-intensity activities, such as exercise or physically demanding tasks. These granular activity data were particularly valuable for understanding the physical demands placed on caregivers and the changing capabilities of patients throughout the recovery process.

#### Mobile Application Data

The Roadmap was used to collect daily self-reported mood scores from both caregivers and patients. The app was developed through an iterative process involving design groups, pilot studies, and extensive literature reviews [14,23,24]. Each day, the app prompted users to rate their mood on a scale of 1–10 at 20:00 everyday during the study period. The participant rates their mood based on a scale of 1–10 [worst possible–best possible]. In addition to mood reporting, the app included multi-component features, such as positive psychology-based activities, health-related resources, and access to physiological data from the Fitbit® device[14]. For participants in the intervention arm, the app provided access to these additional features, while those in the control arm only had access to the mood reporting and Fitbit® data display.

#### PROMIS® Health Measures

The Patient-Reported Outcomes Measurement Information System (PROMIS®)[15] is a collection of person-focused assessments designed to evaluate and track physical, mental, and social health in both adults and children. It was developed and validated with state-of-the-science methods to be psychometrically sound and to transform patient-reported outcome (PRO) assessment[25]. To assess various aspects of health-related quality of life (HRQOL), we administered questionnaires using standardized PROMIS® short forms to both caregivers and patients at different key timepoints: prior to the intervention (baseline), during the intervention (day 30 post-transplant), following the completion of the intervention (120 days post-transplant). The PROMIS® measures used in this study included Global Health[26], emotional distress (Anxiety[27] and Depression[28]), Fatigue[29], Sleep Disturbance[30], Ability to Participate in Social Roles and Activities[31], Companionship[32], Self-Efficacy for Managing Symptoms, and Self-Efficacy for Managing Daily Activities[33].

#### Clinical Events

Clinical data, including transplant-related complications and health care utilization, were collected from the electronic health record (EPIC System, MiChart). Clinical events included: acute graft-versus-host Disease (GVHD), chronic GVHD, infections details (viral, bacterial, fungal), relapse, mortality, total count of hospital days, and readmissions. Graft-versus-Host Disease (GVHD) is a common and potentially serious complication of allogeneic (non-self donor) HCT[34]. GVHD occurs when the donor’s immune cells (the graft) recognize the patient’s tissues (the host) as foreign and attack them. This condition is typically classified into two forms: acute and chronic GVHD[35,36]. By collecting clinical events data along with PROs and physiological data from wearable devices, we aimed to provide a comprehensive view of the HCT experience and the potential correlations of our intervention on both caregivers and patients.

### Data Processing

All data processing was performed using R (version 4.4), primarily utilizing the data.table and dplyr packages for efficient data manipulation.

#### Data Storage

The collected data was stored in three databases. Seven distinct tables from an Oracle database provided the core physiological and behavioral data. These included participant-reported mood entries from the Roadmap app and physiological measurements (heart rate, sleep patterns, and step counts) collected through Fitbit® devices. The PROMIS measures were collected through Qualtrics surveys[37], while clinical outcome was extracted from the electronic health record. Additionally, reference tables that provided device-to-participant mapping, study participant IDs, and demographic information were also exported from Qualtrics.

#### Data Transformation

The raw data was first loaded into R, where all the subsequent processing took place. Initial processing involved filtering out pediatric cases and establishing device-participant relationships through a two-step process: first identifying unique caregiver and patient IDs from the study cohort, then mapping these IDs to their corresponding device identifiers. This phase established the foundation for subsequent data transformation and analysis steps while maintaining all temporal relationships and participant groupings.

Data transformation began with a consistent group assignment process across all datasets. Using a standardized function, we classified each record as either “Patient” or “Caregiver” based on the appropriate identifier–participant IDs for mood,sleep, questionnaire, and clinical outcome data, and device IDs for heart rate and step data. Records not matching either category were excluded from further analysis to maintain data integrity.

The temporal progression of mood scores for both groups across the post-transplant period presented interesting patterns in both magnitude and timing of responses. As shown in Figure 3, not only did the average mood scores vary between caregivers and patients throughout the recovery period, but the distribution of reporting times also revealed distinct behavioral patterns between the two groups, providing insights into their daily routines and emotional reporting habits.

**Figure 3:**
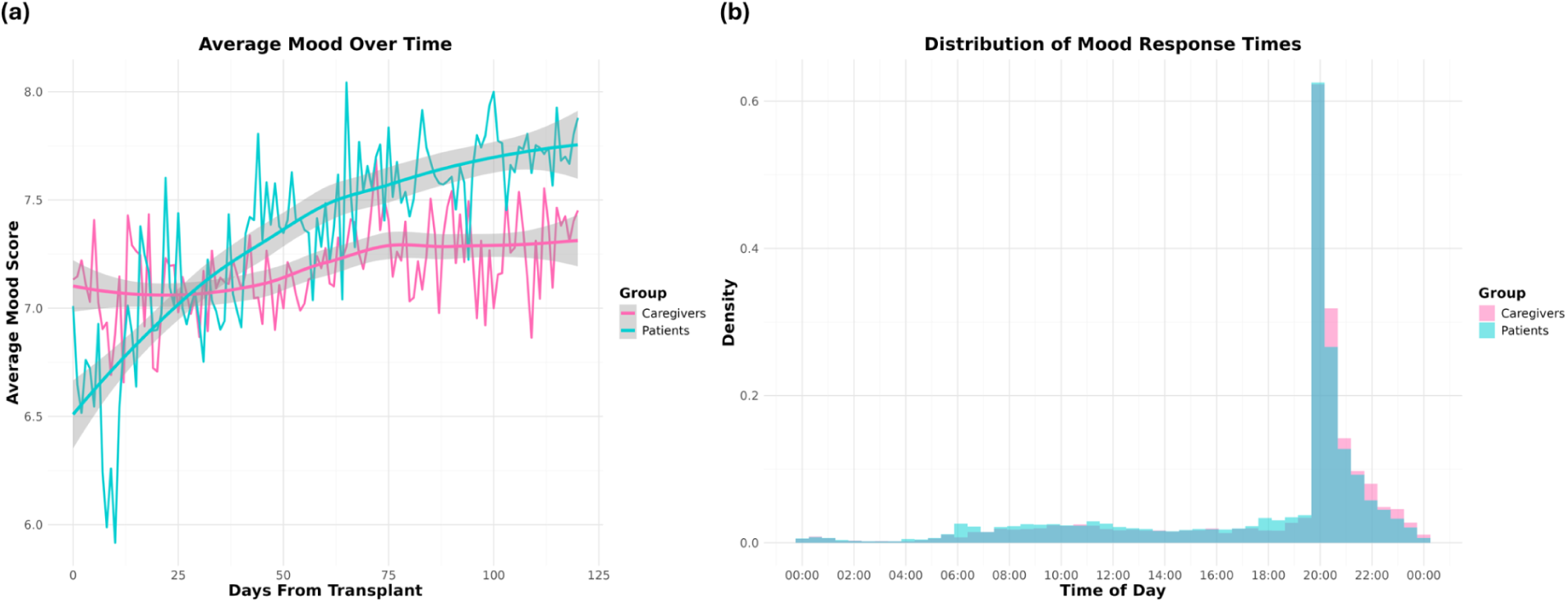
Temporal progression and daily distribution of mood reports in HCT caregiver-patient dyads. Visualization of mood reporting patterns across the 120-day post-transplant period. (A) Average mood scores (± SE) over time for patients (cyan) and caregivers (pink), showing temporal trends in emotional well-being. (B) Distribution of mood reporting times throughout the day. Data represents 20316 mood entries from 157 caregiver-patient dyads.

Sleep data analysis required special consideration due to the presence of two distinct types of sleep measurements. The Fitbit® Charge 3 devices provided both classic sleep metrics and detailed sleep stage data, which were separated and processed separately. Figure 4 presents a comprehensive view of sleep patterns throughout the post-transplant period. The classic metrics Figure 4(a) reveal overall sleep quality indicators such as total sleep time and efficiency, while the sleep stage analysis Figure 4(b) provides deeper insights into the architecture of sleep for both caregivers and patients.

**Figure 4:**
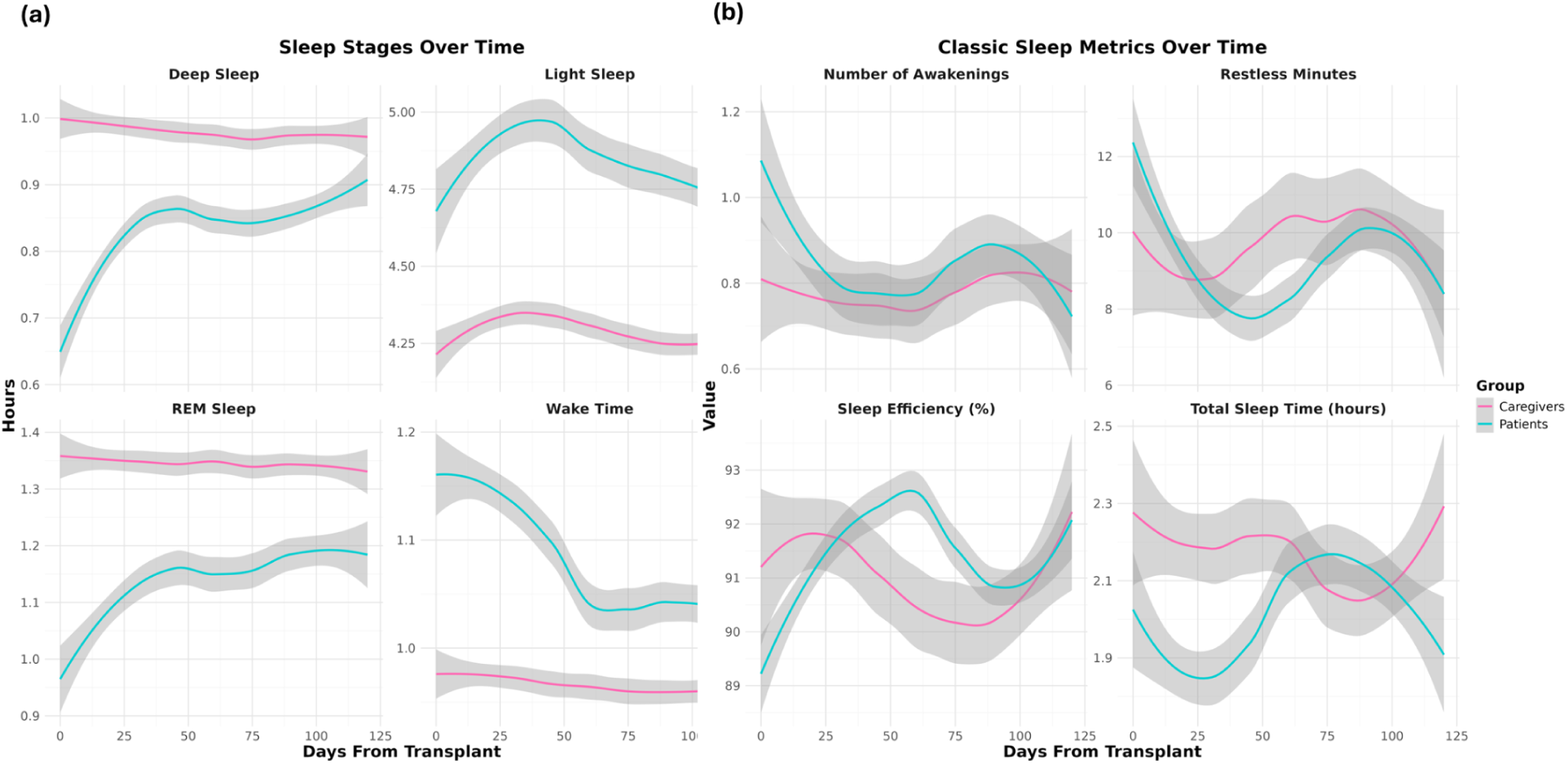
Temporal progression of classic sleep metrics in HCT caregiver-patient dyads. (a) Classic sleep metrics showing LOESS smoothed trends (± SE) for total sleep time (hours), sleep efficiency (%), number of awakenings, and restless minutes. (b) Stages sleep metrics showing LOESS smoothed trends (± SE) for time spent in deep sleep, light sleep, REM Sleep, and wake time (hours). All metrics derived from Fitbit® Charge 3 devices, with curves representing averaged values across all participants in each group.

The continuous physiological monitoring via Fitbit® devices generated massive datasets required careful processing and validation. As shown in Figure 3, the distribution of these measurements revealed distinct patterns between caregivers and patients. Heart rate data, comprising over 5,000,000 individual measurements at minute-level granularity, was carefully synchronized with device activation periods to account for participants who received multiple Fitbit devices during the study. The step count data presented a particular challenge, with over 450,000,000 initial records including numerous invalid entries from sedentary activity logging. To ensure data quality, we used the more reliable heart rate data timestamps as a filtering criterion for step counts, effectively removing spurious entries while preserving genuine physical activity records. The resulting distributions are presented in Figure 5

**Figure 5:**
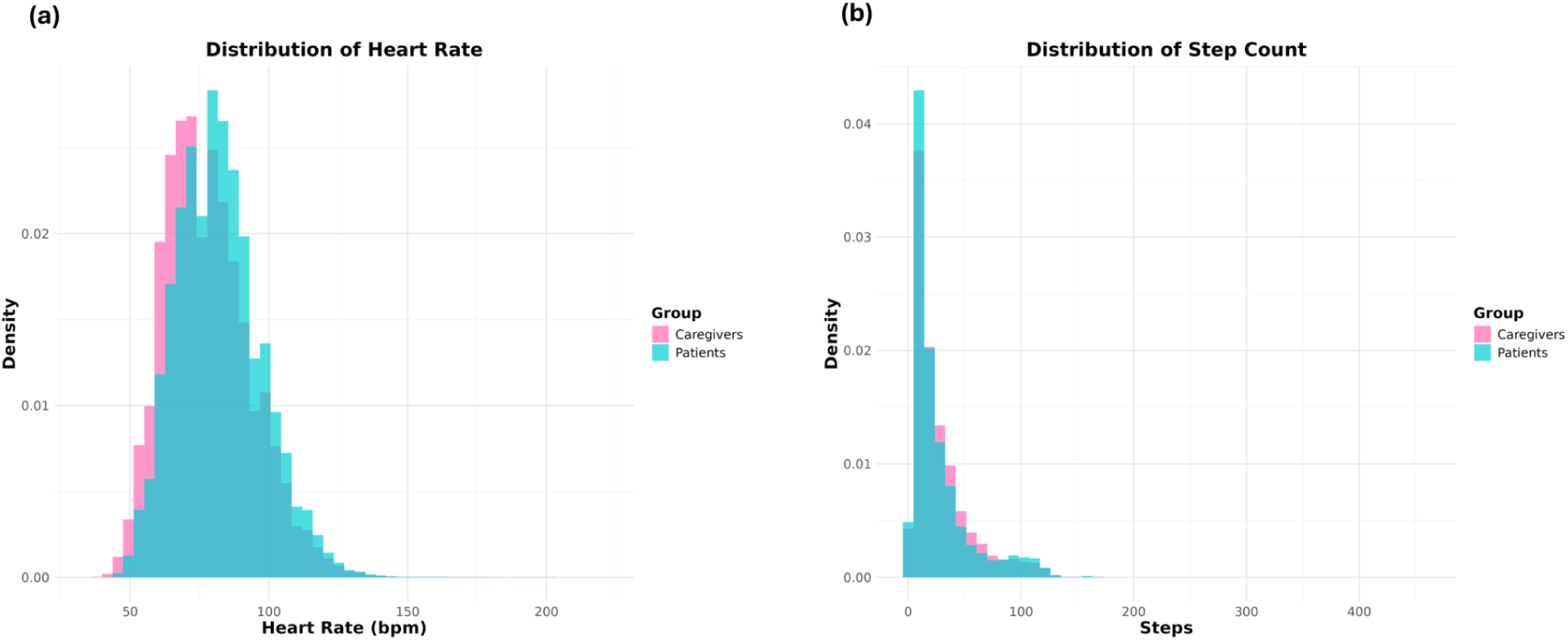
Histogram of physiological measurements in dCT caregiver-patient dyads. (a) Heart Rate. (b) Number of steps taken per minute

#### PROMIS® T-Score Calculation

For the PROMIS® health measures, we utilized the HealthMeasures Scoring Service [35] to process both caregiver and patient responses for each PROMIS® instrument. This scoring method employed “response pattern scoring,” which accounted for the responses to each item for every participant. Response pattern scoring was particularly advantageous when dealing with missing data, such as when a participant skipped an item or when different groups answered different sets of items. After scoring the responses, we converted the raw scores into a T-score metric [36], a standardized score that represents an individual’s health status or function relative to a reference population. To illustrate the distribution of the processed T-score data, Figure 6 displays one of the PROMIS® health measures (Anxiety), for both caregivers and patients at different time points (Baseline, Day 30, Day 120 post-HCT). This visualization highlighted the differences in anxiety levels across caregivers and patients and time periods, providing a graphical display of the data after processing.

**Figure 6:**
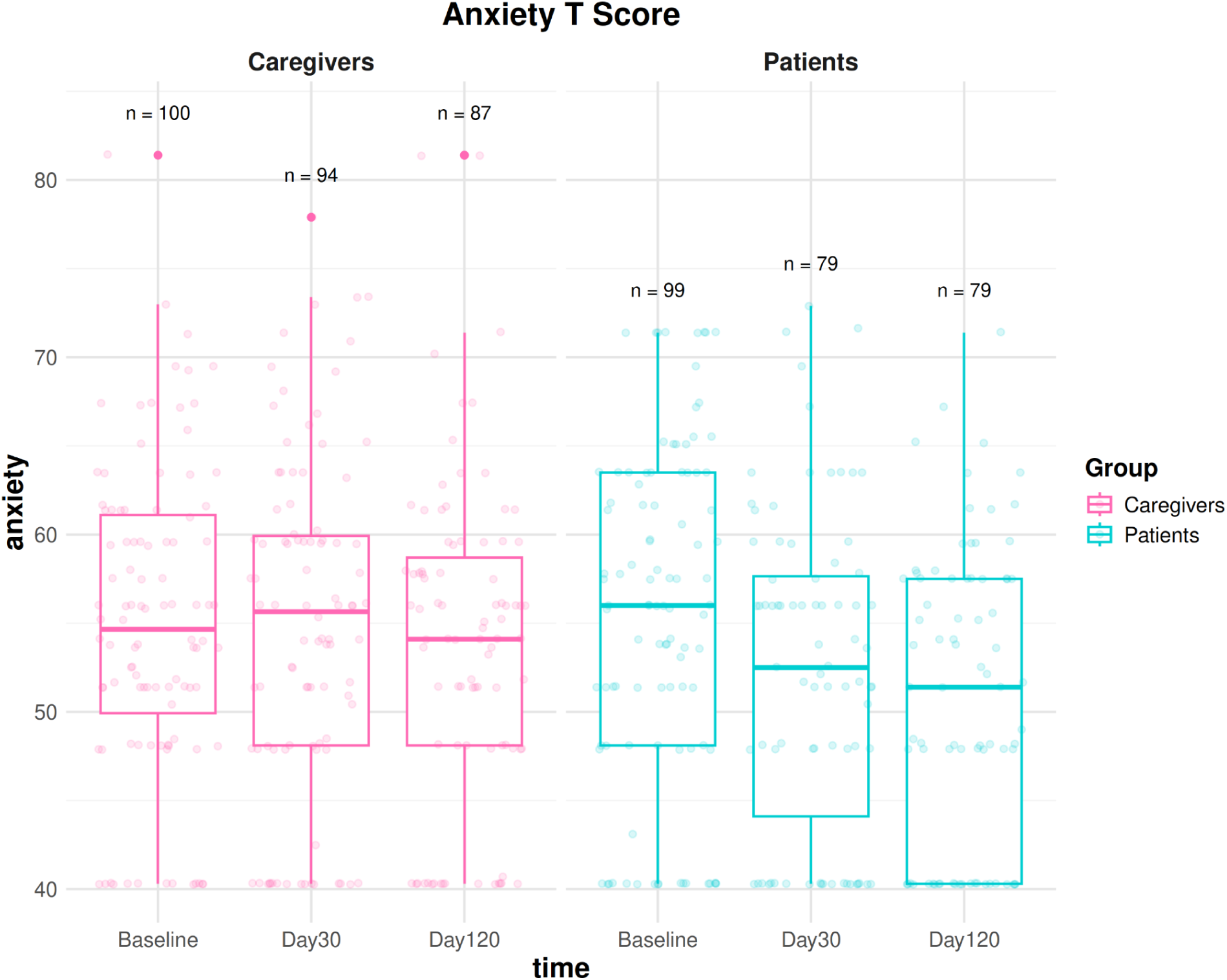
Longitudinal PROMIS® Anxiety T-Scores in HCT caregiver-patient dyads. Distribution of PROMIS® Anxiety T-scores at three key timepoints during the post-transplant period: Baseline, Day 30, and Day 120. Box plots show median, quartiles, and outliers for both patients (cyan) and caregivers (pink), with individual data points overlaid to show the complete distribution. T-scores are standardized with a population mean of 50 and standard deviation of 10, where higher scores indicate greater anxiety. Data processed using HealthMeasures Scoring Service with response pattern scoring to account for any missing responses.

#### Clinical Events Extraction

Clinical event data were filtered to include only events occurring after each patient’s transplant date. The final datasets consist of three tables: infections, readmissions, and clinical outcomes (relapse, GVHD, death). Figure 7 illustrates the distribution of major clinical outcomes in our patient cohort through a stacked bar plot. The visualization captures four critical events: severe acute GVHD (grades 3-4), chronic GVHD, death, and disease relapse. A notable feature of the data is the presence of missing values (NA) in both GVHD categories, which reflects an important clinical distinction in our cohort. These missing values correspond to patients who underwent autologous transplants, where their own stem cells were used for the procedure. As these patients received their own cells rather than donor cells, they do not face the risk of GVHD, making these measurements inapplicable to their cases[38].

**Figure 7:**
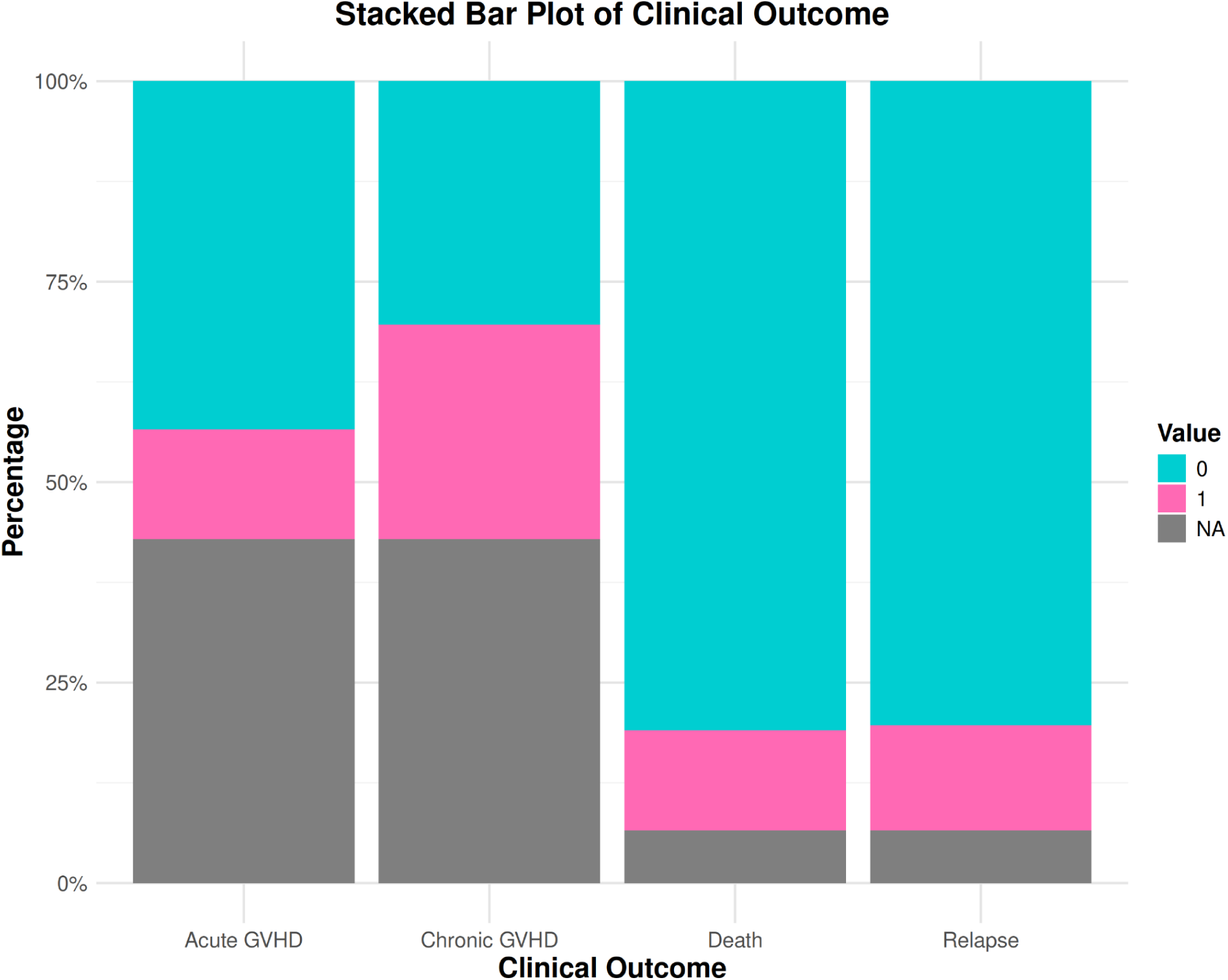
Distribution of major clinical outcomes in HCT recipients. Stacked bar plot showing the occurrence of key clinical outcomes in the study cohort. Outcomes include acute Graft-versus-Host Disease (grades 3-4), chronic GVHD, death, and disease relapse. Gray bars (NA) in GVHD categories represent autologous transplant recipients, for whom GVHD is not applicable due to the use of their own stem cells. Values show the percentage of the total cohort for each outcome.

Moving to the readmission and infection data, data tables were structured at the event level, describing each readmission and infection occurrence. Figure 8 shows a histogram of two key variables: the number of readmissions and the number of infections, aggregated for each patient. The blue bars indicate the number of infections per patient, while the pink bars represent the number of readmissions per patient. The majority of patients experienced no readmissions or infections, reflected by the highest count at zero events. As the number of events increased, the frequency of patients with multiple readmissions or infections decreased.

**Figure 8:**
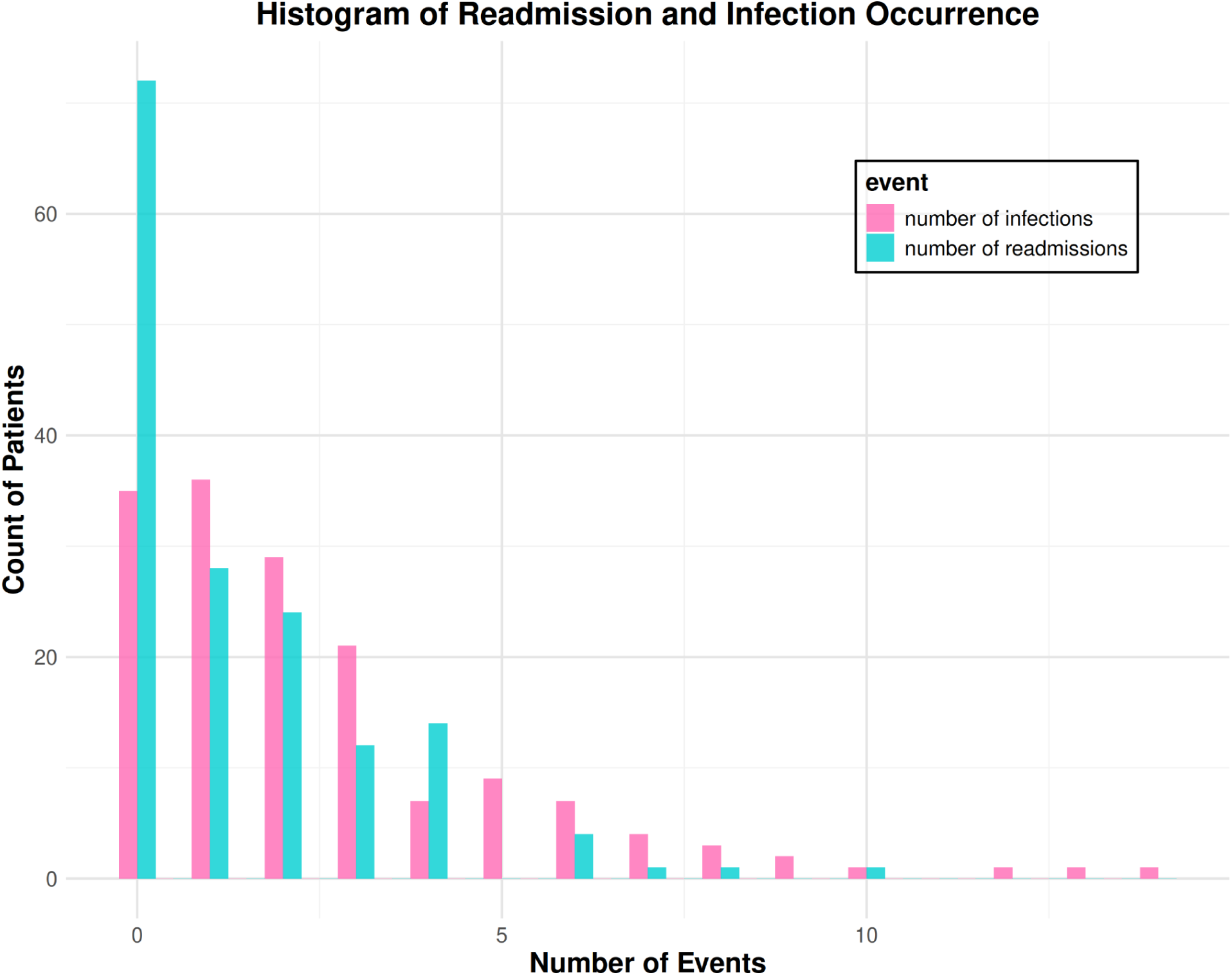
Distribution of readmissions and infections in HCT recipients. Frequency distribution of post-transplant complications per patient. (A) Number of recorded infections per patient (blue). (B) Number of hospital readmissions per patient (pink). Both distributions show right-skewed patterns with the majority of patients experiencing zero or few events, and decreasing frequencies for higher event counts. Y-axis represents the count of patients, while X-axis shows the number of events (readmissions or infections) experienced. Note the different scales for readmissions and infections, reflecting their distinct occurrence patterns in the post-HCT period

#### Data Censoring

Privacy protection centered around a comprehensive ID anonymization process. We developed a preprocessing function that created anonymous identifiers for both caregiver and patient information. These anonymous identifiers replaced participant IDs with anonymous participant codes (e.g., “P001”), and device IDs linked to participants with anonymous device codes (e.g., “P001_D01”).

The censoring followed a systematic approach across all datasets while accommodating their unique characteristics. The processing steps included calculating the number of days between each timestamp and the transplant date, rather than using the exact event date. All the wearable data retained scores and relative timing while the survey metadata and participant identifiers.

#### Data Aggregation

The final phase of processing created daily summaries of high resolution physiological measurements, which included activity, heart rate, and step data, to facilitate longitudinal analysis while maintaining manageable data volumes. Heart rate summaries captured both basic statistical measures and circadian patterns. The basic statistics included central tendency measures (mean, median), variability measures (standard deviation, IQR), and range statistics (minimum, maximum, quartiles) (Figure 9a). Circadian metrics provided insight into daily rhythms through time-based averages and specific rhythm parameters such as amplitude, mesor, acrophase, and bathyphase (Figure 9d).

**Figure 9:**
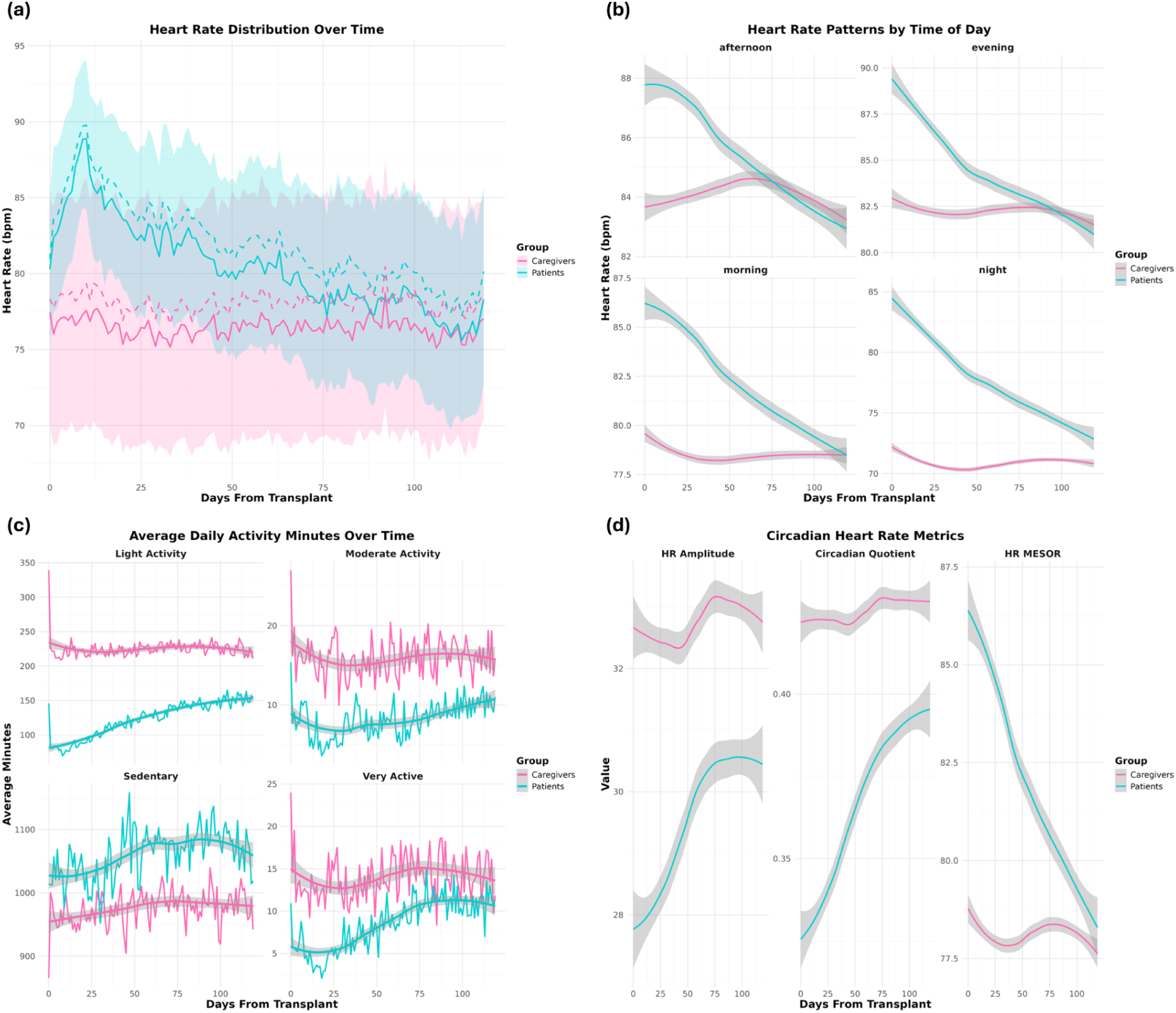
Temporal patterns of heart rate metrics in HCT caregiver-patient dyads. (A) Daily heart rate distributions showing median (solid lines) and mean (dashed lines) values with interquartile ranges (shaded areas) for patients (cyan) and caregivers (pink). (B) Heart rate patterns stratified by time of day (morning: 6:00-12:00, afternoon: 12:00-18:00, evening: 18:00-24:00, night: 00:00-6:00), displaying LOESS smoothed trends with 95% confidence intervals. Data derived from minute-level Fitbit® measurements aggregated daily. (C) Distribution of daily activity levels showing average minutes spent in sedentary, light, moderate, and very active states. All panels show LOESS smoothed trends with 95% confidence intervals. Activity classifications derived from Fitbit® intensity measures based on movement and heart rate patterns..(D) Temporal progression of key circadian metrics: HR Amplitude (peak-to-trough difference), MESOR (rhythm-adjusted mean), and Circadian Quotient (amplitude/MESOR ratio).

Activity summaries were generated using both step counts and activity classifications. Step metrics included daily totals and their temporal distribution, while activity classifications provided detailed breakdowns of time spent at different activity levels. The summary incorporated coverage statistics and data completeness indicators to ensure quality control in subsequent analyses. Figure 9c displays the temporal progression of different activity types for both caregivers and patients throughout the study period.

Throughout the temporal aggregation process, we maintained group classifications and transplant timeline references while ensuring computational efficiency. The final processed datasets were exported as CSV files, featuring consistent structure and anonymized identifiers while preserving the analytical utility necessary for investigating physiological and behavioral patterns in HCT patients and their caregivers.

### Data Validation

To ensure the reliability and quality of our dataset, we implemented a comprehensive validation framework encompassing multiple dimensions of data quality assessment. We analyzed data completeness, timing patterns, and adherence to expected measurement protocols across both caregiver and patient groups.

#### Fitbit Compliance

**Figure 10:**
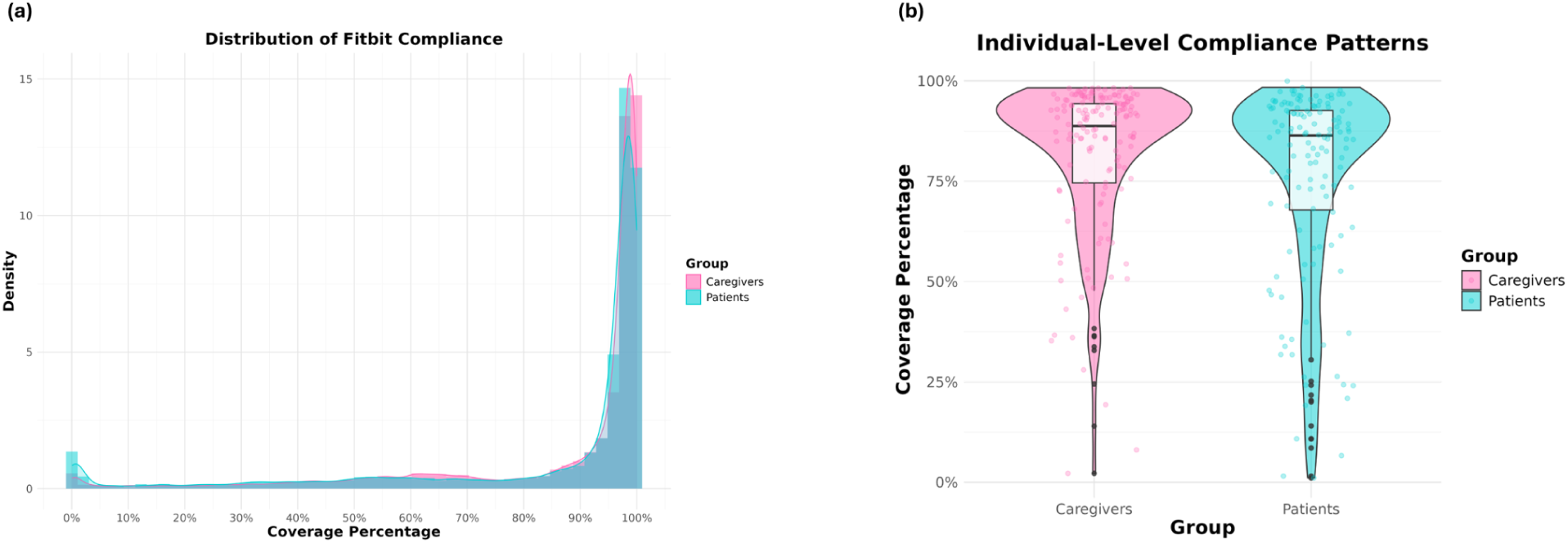
Fitbit® data completeness and compliance patterns. Analysis of Fitbit® data completeness in HCT caregiver-patient dyads. (A) Distribution of daily heart rate data coverage showing median (patients: 97.3%, caregivers: 97.7%) and mean (patients: 84.0%, caregivers: 86.4%) coverage rates with standard deviations. (B) Individual-level compliance patterns showing the distribution of participant-specific coverage rates (box plots) and day-to-day variability (violin plots) for both groups. Patient coverage shows greater variability (SD = 24.5%) compared to caregivers (SD = 18.4%). Early post-transplant period (first 30 days) highlighted to show distinct patterns in monitoring adherence

#### Overall Compliance

The analysis of heart rate data coverage revealed distinct patterns in monitoring compliance (Figure 10). While median daily coverage rates were excellent for both patients (97.3%) and caregivers (97.7%), there was considerable variability in the data. Mean coverage rates were somewhat lower than medians (84.0% for patients and 86.4% for caregivers), with substantial standard deviations (26.5% and 22.8% respectively), indicating positively skewed distributions with some low-coverage days pulling down the averages.

#### Participant-Level Variation

Individual-level analysis revealed important patterns in monitoring adherence. Caregivers showed higher average compliance (81.4%) compared to patients (75.4%). Patient coverage was more variable (SD = 24.5%) than caregiver coverage (SD = 18.4%). Individual compliance rates ranged widely from 2.2% to 98.3% for caregivers and 1.1% to 98.4% for patients. The median participant-level compliance was better than average (86.4% for patients, 88.7% for caregivers), again indicating some participants with particularly low coverage affecting group averages.

Among caregivers, 25.3% demonstrated highly consistent compliance (day-to-day standard deviation < 10%). While the corresponding statistic for patients was 13.5%

#### Early Post-Transplant Period

The early post-transplant period (first 30 days) showed distinct patterns with higher variability among patients (coefficient of variation = 36.9%) compared to caregivers (25.6%). A lower mean compliance for patients (80.6%) compared to caregivers (86.5%) was present along with a greater standard deviation in daily coverage for patients (29.8%) compared to caregivers (22.2%)

These patterns suggest that the immediate post-transplant period may be particularly challenging for maintaining consistent device wear, especially for patients.

These findings indicate that while overall compliance was good, there were important differences between caregivers and patients, and considerable variation among individuals. The early post-transplant period appears to be a particularly critical time for monitoring compliance, especially among patients.

#### Mood Response

##### Response patterns

Analysis of mood response data revealed distinct patterns in data collection compliance. The distribution of response times showed clear temporal preferences, with evening hours (17:00-22:00) being the most common reporting time for both patients (66.8%) and caregivers (68.6%). Morning responses (5:00-12:00) accounted for 15.3% of patient responses and 12.4% of caregiver responses, while afternoon (12:00-17:00) and night (22:00-5:00) periods showed lower reporting frequencies (patients: 9.1% afternoon, 8.7% night; caregivers: 8.5% afternoon, 10.6% night). This suggests that the reminder prompt displayed on the app at 20:00 most likely dictates the time that the participants report their mood

##### Response Frequency

Daily reporting patterns indicated consistent participation, with patients averaging 1.22 (SD = 0.49) reports per day and caregivers averaging 1.14 (SD = 0.44) reports per day. The majority of days featured single reports (80.2% for patients, 87.8% for caregivers), with the remaining days showing multiple reports. Notably, there were no days with zero reports recorded, suggesting good overall engagement with the mood reporting protocol.

##### Missing data patterns

The analysis revealed several key patterns in missing data. Patients experienced an average of 12.5 gaps in reporting throughout the study period, while caregivers averaged 15.7 gaps. The maximum gap length was 83 days for patients and 89 days for caregivers. The median gap length was 1 day for both groups, though mean gap lengths were longer (5.83 days for patients, 4.31 days for caregivers). On average, patients missed 39.9 days of reporting, while caregivers missed 36.8 days.

##### Sleep Measurement

**Figure 11:**
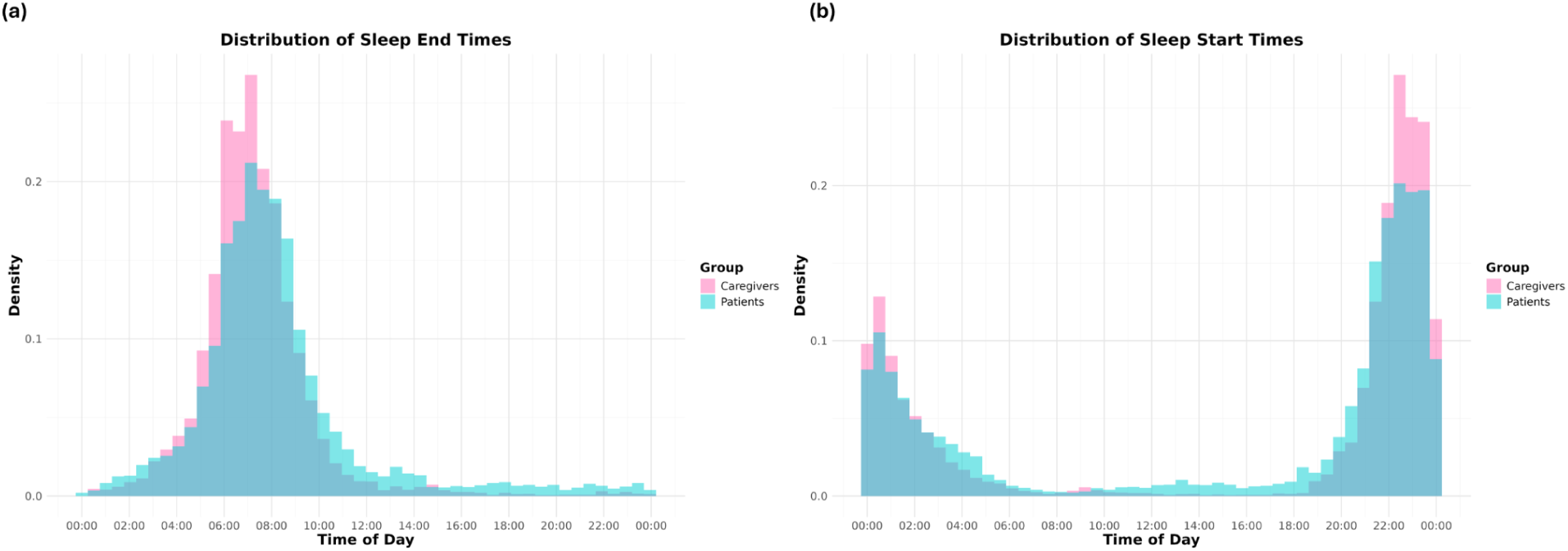
Distribution of sleep onset and wake times. (a) Distribution of sleep onset (mode: patients 22:28, caregivers 22:30) and wake times (median: patients 07:16, caregivers 07:03). (b) Longitudinal analysis of sleep timing compliance showing stable patterns with higher adherence in caregivers (88.3%) compared to patients (77.0%) Use the mode instead of the median.

##### Sleep Data Validation

Sleep timing analysis (Figure 11) revealed generally appropriate nocturnal sleep patterns, with most sleep episodes occurring during expected nighttime hours. The median sleep onset time was 22:28 for patients and 22:30 for caregivers, while median wake times were 07:37 and 07:03 respectively. This pattern suggests good compliance with normal sleep-wake cycles and validates the reliability of the sleep measurements.

##### Sleep compliance

Sleep timing compliance defined as sleep onset between 20:00-04:00 and wake time between 04:00-12:00, showed high adherence rates. Caregivers demonstrated higher compliance rates (88.3%) compared to patients (77.0%). This difference was consistent across both sleep onset (93.5% vs 85.1% compliance) and wake times (91.8% vs 84.2% compliance). The longitudinal analysis demonstrates that this pattern remained relatively stable throughout the study period, though with some variation in compliance rates over time.

#### Survey measures

##### PROMIS® T-score Correlation

Figure 12 shows a heatmap of the correlation among PROMIS® T-scores. Positive health measures exhibited strong positive correlations with one another, while negative measures displayed strong negative correlations with positive measures. This trend supported the internal consistency of the data, reinforcing its validity and the expected interrelationships between physical, mental, and cognitive health metrics.

**Figure 12:**
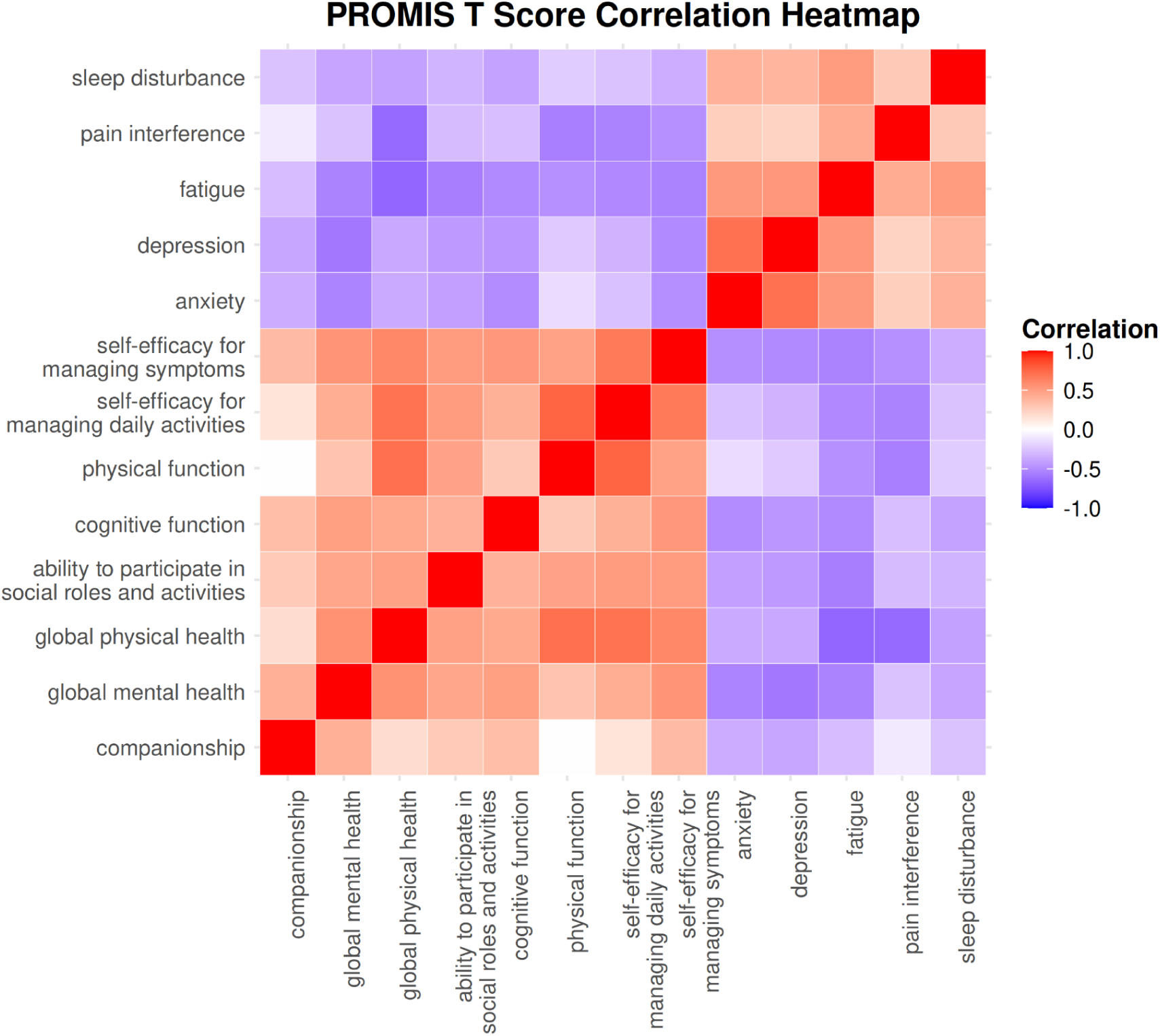
Correlation heatmap for PROMIS® T-scores. Positive PROMIS® measures includes Companionship, Global Mental Health, Global Physical Health, Ability to Participate in Social Roles and Activities, Cognitive Function, Physical Function, Self-Efficacy for Managing Daily Activities, and Self-Efficacy for Managing Symptoms, are indicators of better health outcomes. In contrast, negative PROMIS® measures, Anxiety, Depression, Fatigue, Pain Interference, and Sleep Disturbance, represent poorer health outcomes.

##### Clinical Events Validation

The clinical events pattern from Figure 13 shows a cluster of culture draws early in the timeline, followed by admissions and more critical outcomes such as GVHD, relapse, and death, reflecting the expected progression of care and disease in this patient group. Notably, no events are recorded after the occurrence of a mortality event (denoted by red triangles), further supporting the validity of the dataset. This logical consistency—where no medical events, such as GvHD or relapse, are flagged after death—reinforces the accuracy of the data, ensuring that it accurately captures patient progress.

**Figure 13:**
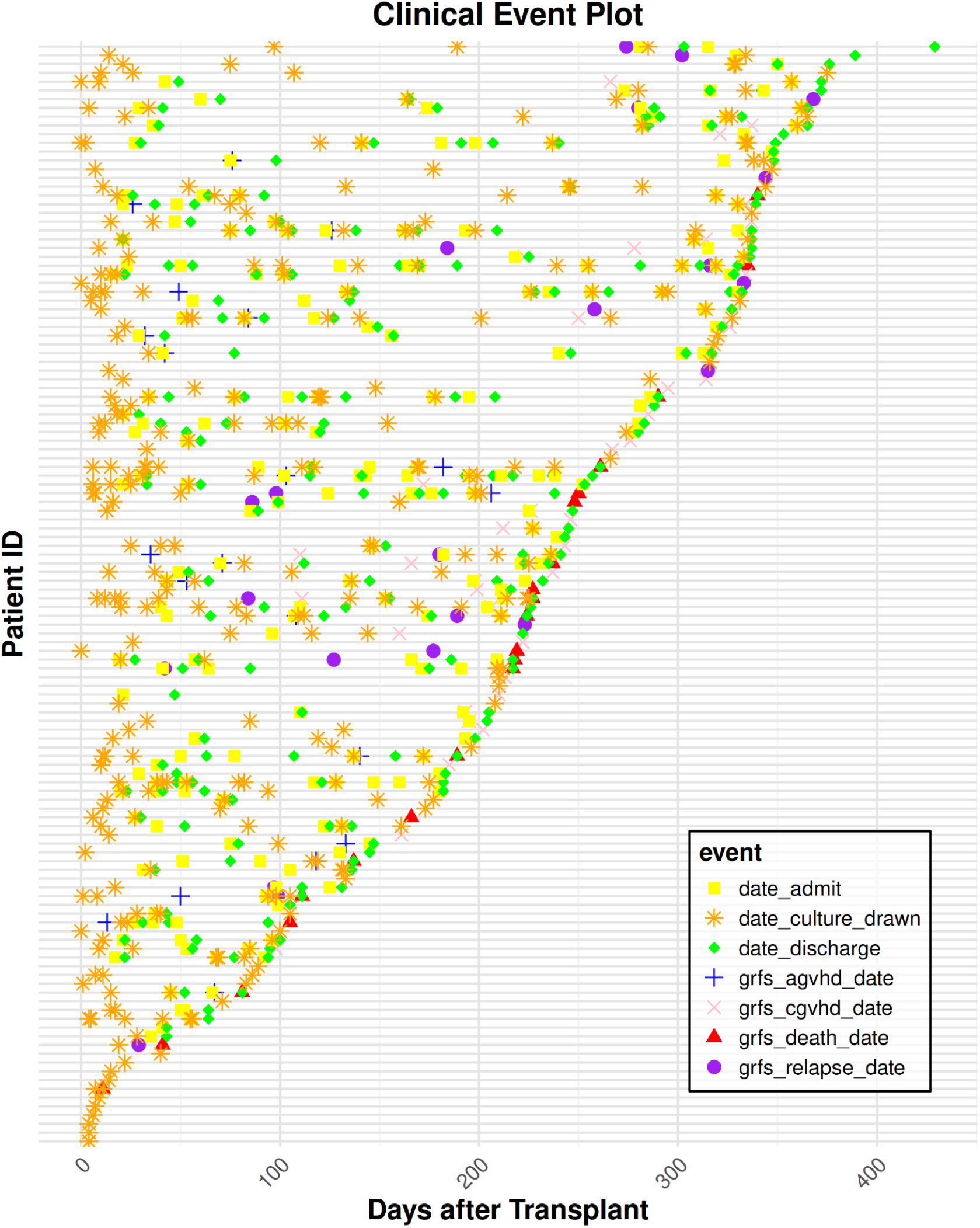
Clinical Events Pattern for each Patient. Events in the daily timeline, starting from the transplant date, include admission dates (yellow squares), dates when culture samples were drawn (orange asterisks), discharge dates (blue crosses), graft-versus-host disease (GvHD) occurrences (acute: green diamonds, chronic: pink X), relapse (purple circles), and mortality (red triangle).

### Reuse Potential

This comprehensive dataset offers significant potential for reuse in various research contexts:

1. **Predictive Modeling:** The continuous physiological data, combined with psychological measures and clinical outcomes, provide a robust foundation for developing predictive models. These models could forecast post-transplant complications, recovery trajectories, or clinical events such as GVHD in HCT patients.
2. **Caregiver/Dyadic Support Interventions**: By analyzing the well-being data of caregivers, this dataset could inform the beneficial effect of interventions aimed at improving caregiver resilience, reducing burden, and enhancing quality of life. Furthermore, dyadic analyses could indicate the benefit of interventions that address the needs of both patients and their caregivers simultaneously.
3. **Circadian Rhythm Studies:** The granular heart rate and activity data allow for detailed investigations into circadian rhythm disruptions in critically ill patients and their caregivers. This information could be instrumental in understanding the impact of HCT and caregiving responsibilities on biological rhythms.
4. **mHealth Intervention Design:** Usage patterns of the Roadmap app can provide critical insights into user engagement and adherence, informing the design of future mHealth interventions. These insights can contribute to more effective and personalized digital health tools for caregiver-patient dyads in a variety of medical contexts.
5. **Dyadic Analyses:** The availability of paired caregiver-patient data offers a unique opportunity for dyadic analysis, exploring how the physical and psychological well-being of caregivers and patients influence one another. Longitudinal assessments could reveal dynamic interactions between the two and help identify critical periods of interdependence.
6. **Stress and Recovery Patterns:** The physiological data, such as heart rate variability and activity levels, may be used to identify patterns of stress and recovery in both caregivers and patients during the HCT process. This could advance understanding of how stress is managed and alleviated across the post-transplant period.
7. **Physical Activity and Sleep Research:** The detailed step and sleep data present opportunities for research into physical disability and sleep disturbances commonly experienced in medical settings. This data could help elucidate the impact of chronic stress, illness, and caregiving on both physical activity levels and sleep quality, contributing to a better understanding of recovery and overall well-being.

**Table 1:** Demographic Data Variables. (N = 334 observations)

| Variable Name | Description | Type | Values |
| --- | --- | --- | --- |
| STUDY_PRTCPT_ID | Anonymous participant identifier | Character | Format: "P####" |
| arm | Study arm assignment | Character | "intervention", "Control" |
| cg_hours | Caregiver weekly commitment | Character | "less or equal to 40 hours", "more than 40 hours" |
| age | Age category | Character | "18-39", "40-60", "61+" |
| gender | Participant gender | Character | "Male", "Female" |
| monthly_income | Monthly household income category | Character | "< \$1,000", "\$1,000 - \$2,999", "\$3,000 - \$4,999", "\$5,000 - \$6,999", "≥ \$7,000" |
| transplant_type | Type of transplant | Character | "Auto" = autologous, "Allo" = allogeneic |
| dyad_id | Unique identifier for caregiver-patient pair | Integer | 1, 2, 3, ... |
| role | Participant role | Character | "Patients", "Caregivers" |
| in_hospital_days | Number of days spent in hospital | Character | Numeric values or "NA - OHSU" |

**Table 2:** Mood Data Variables. (N = 20,316 observations)

| Variable Name | Description | Type | Range/Values |
| --- | --- | --- | --- |
| STUDY_PRTCPT_ID | Anonymous participant identifier | Character | Format: "P####" |
| MOOD | Self-reported mood score | Integer | 1-10 |
| Group | Participant category | Character | "Patients",<br>"Caregivers" |
| time_stamp | Time of mood report | Character | 24-hour format<br>(HH:MM:SS) |
| DaysFromTransplant | Days elapsed since transplant | Integer | 0-120 |

**Table 3:** Sleep Data Variables - Classic Method. (N = 8,639 observations)

| Variable Name | Description | Type | Units |
| --- | --- | --- | --- |
| STUDY_PRTCPT_ID | Anonymous participant identifier | Character | Format: "P####" |
| ASLEEP_VALUE | Total time asleep | Integer | Minutes |
| INBED_VALUE | Total time in bed | Integer | Minutes |
| ASLEEP_MIN | Minutes asleep | Integer | Minutes |
| ASLEEP_COUNT | Number of sleep episodes | Integer | Count |
| AWAKE_COUNT | Number of awake episodes | Integer | Count |
| AWAKE_MIN | Minutes awake | Integer | Minutes |
| RESTLESS_COUNT | Number of restless episodes | Integer | Count |
| RESTLESS_MIN | Minutes restless | Integer | Minutes |
| TYPE | Sleep tracking method | Character | "classic" |
| Group | Participant category | Character | "Patients",<br>"Caregivers" |
| DaysFromTransplant | Days elapsed since transplant | Integer | 0-120 |
| SLEEP_START_TIME | Sleep start time | Character | 24-hour format (HH:MM:SS) |
| SLEEP_END_TIME | Sleep end time | Character | 24-hour format (HH:MM:SS) |

**Table 4:**
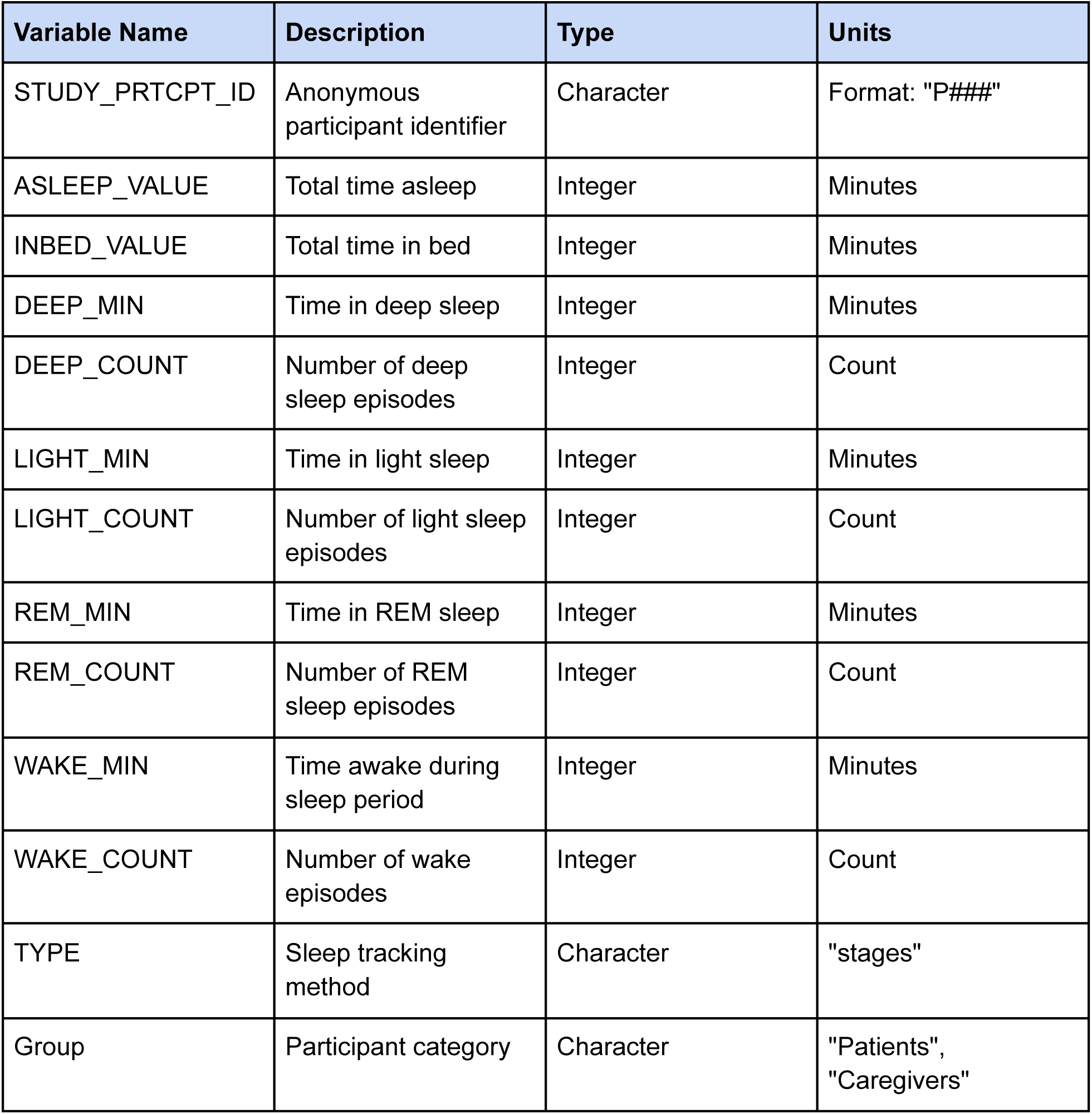
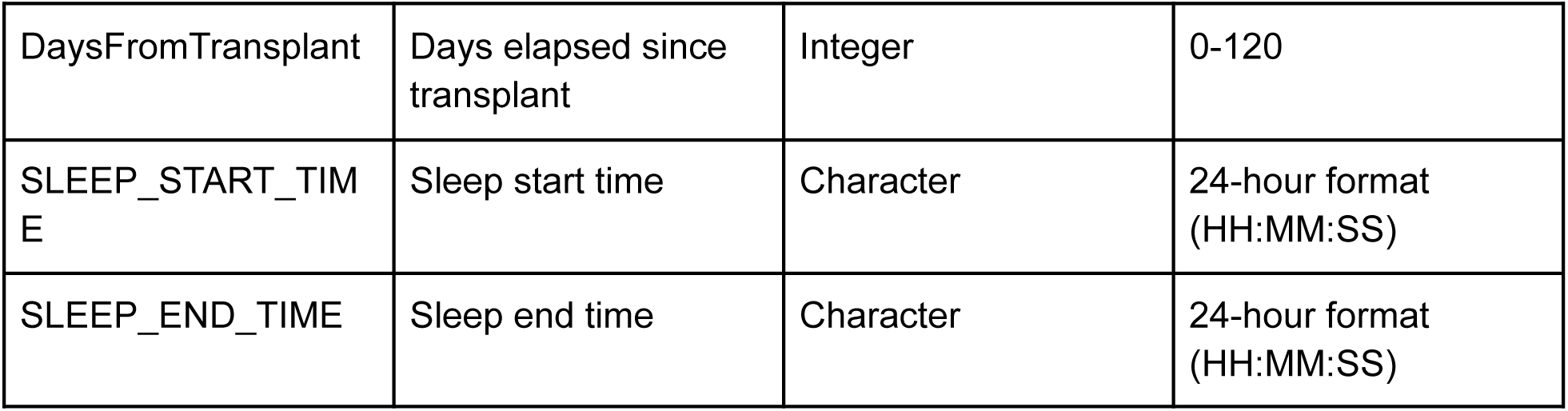
Sleep Data Variables - Stages Method. (N = 20,136 observations)

**Table 5:**
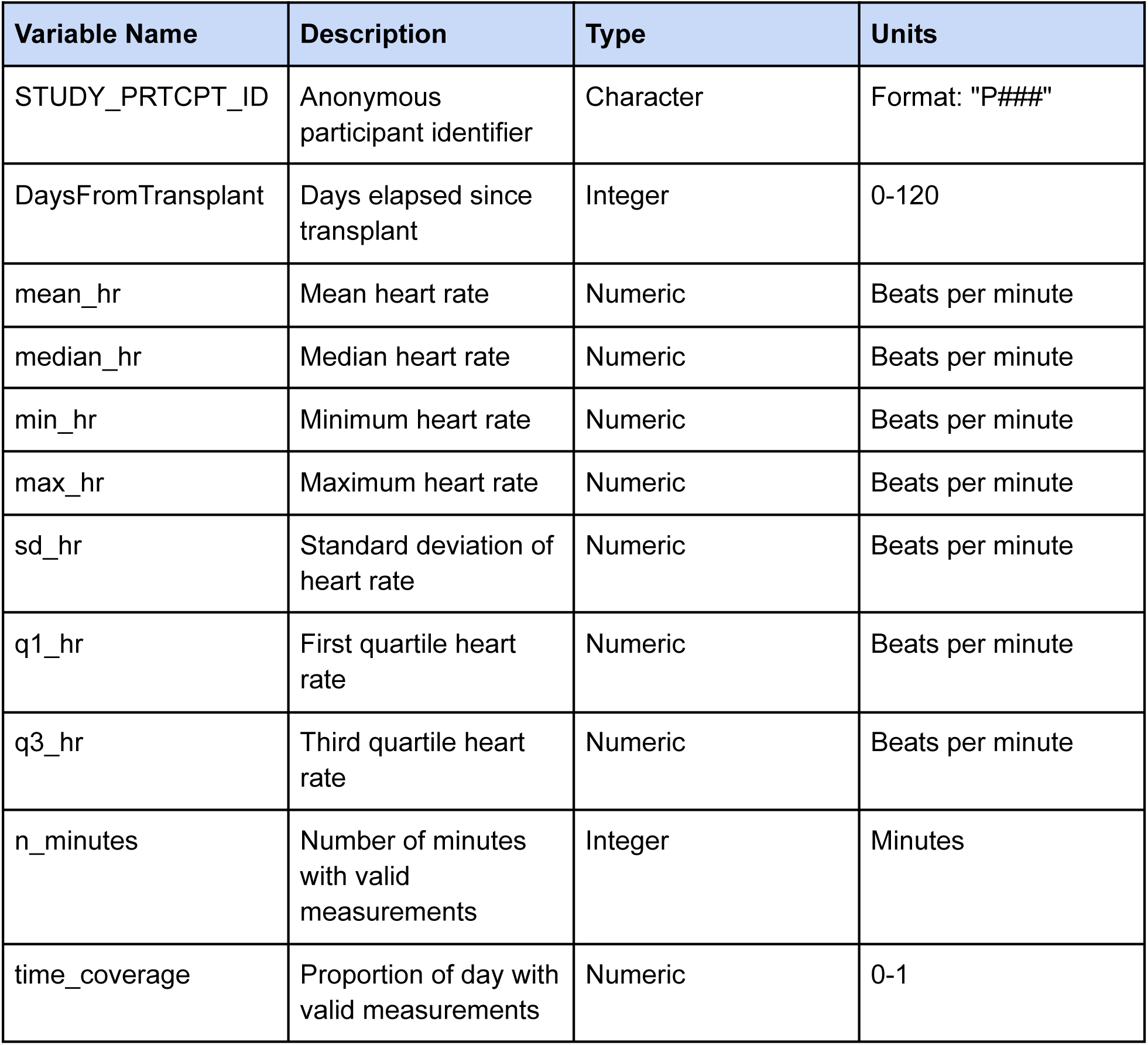
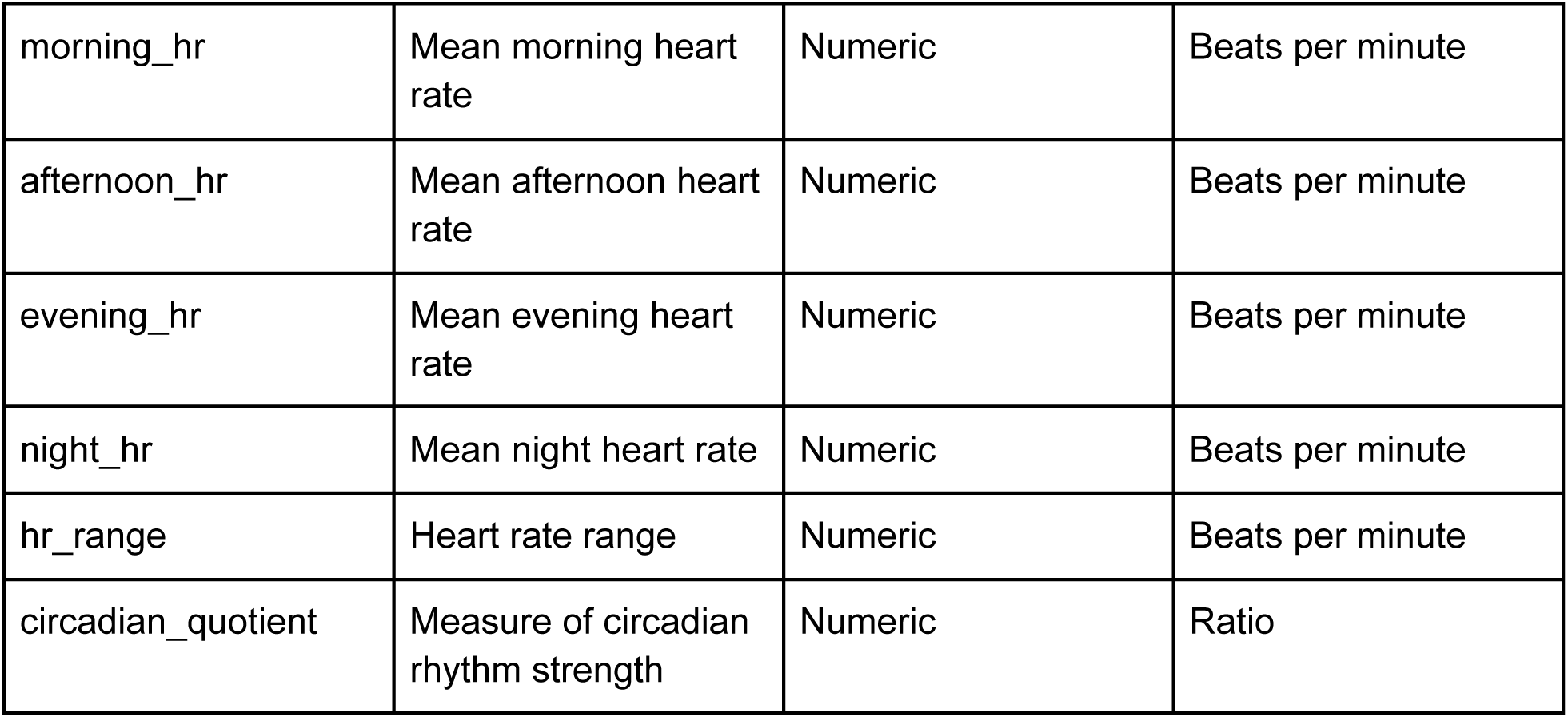
Daily Heart Rate Summary Variables. (N = 24,587 observations)

| Variable Name | Description | Type | Units |
| --- | --- | --- | --- |
| STUDY_PRTCPT_ID | Anonymous participant identifier | Character | Format: "P####" |
| DaysFromTransplant | Days elapsed since transplant | Integer | 0-120 |
| mean_hr | Mean heart rate | Numeric | Beats per minute |
| median_hr | Median heart rate | Numeric | Beats per minute |
| min_hr | Minimum heart rate | Numeric | Beats per minute |
| max_hr | Maximum heart rate | Numeric | Beats per minute |
| sd_hr | Standard deviation of heart rate | Numeric | Beats per minute |
| q1_hr | First quartile heart rate | Numeric | Beats per minute |
| q3_hr | Third quartile heart rate | Numeric | Beats per minute |
| n_minutes | Number of minutes with valid measurements | Integer | Minutes |
| time_coverage | Proportion of day with valid measurements | Numeric | 0-1 |
| morning_hr | Mean morning heart rate | Numeric | Beats per minute |
| afternoon_hr | Mean afternoon heart rate | Numeric | Beats per minute |
| evening_hr | Mean evening heart rate | Numeric | Beats per minute |
| night_hr | Mean night heart rate | Numeric | Beats per minute |
| hr_range | Heart rate range | Numeric | Beats per minute |
| circadian_quotient | Measure of circadian rhythm strength | Numeric | Ratio |

**Table 6:**
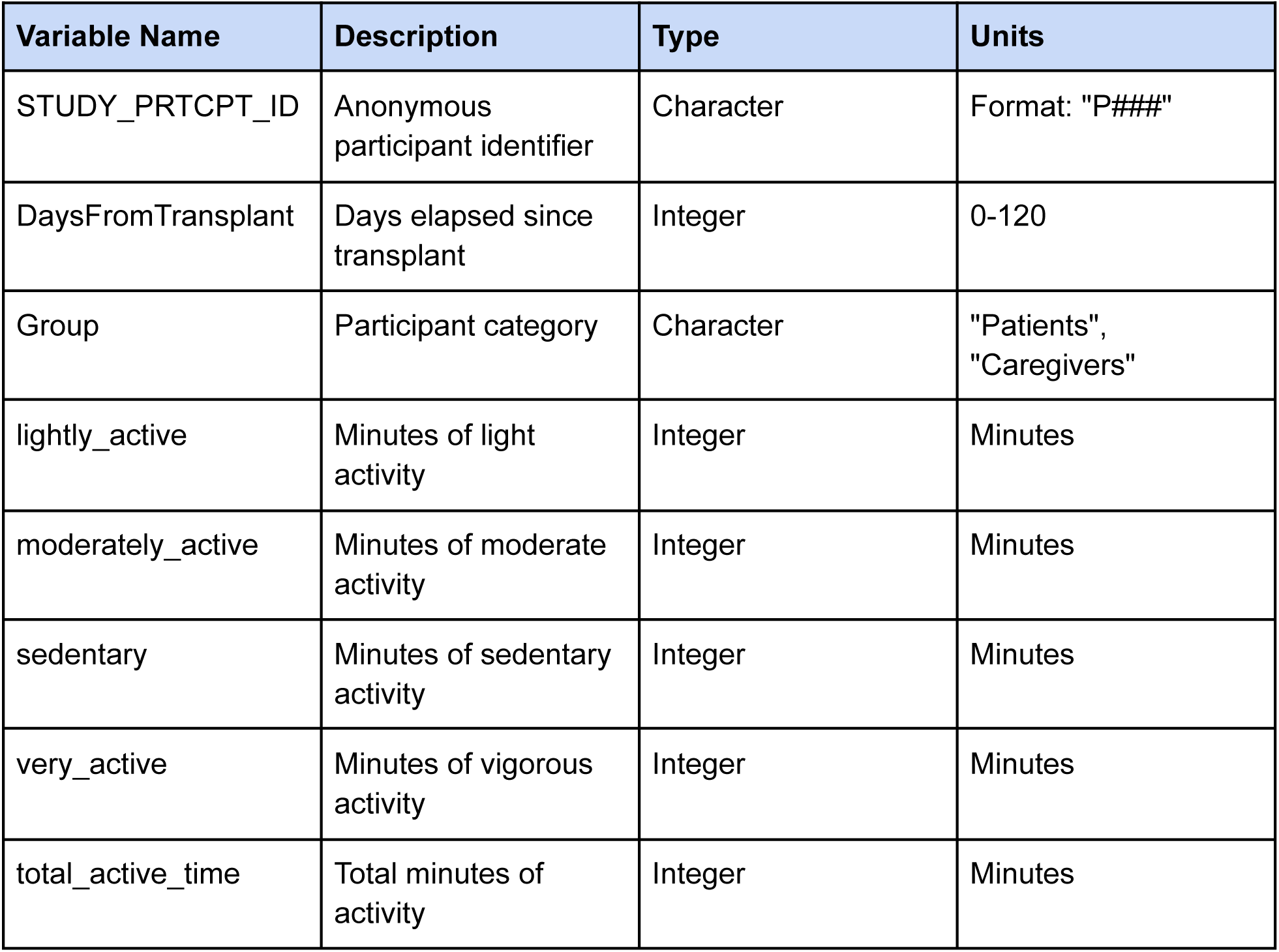
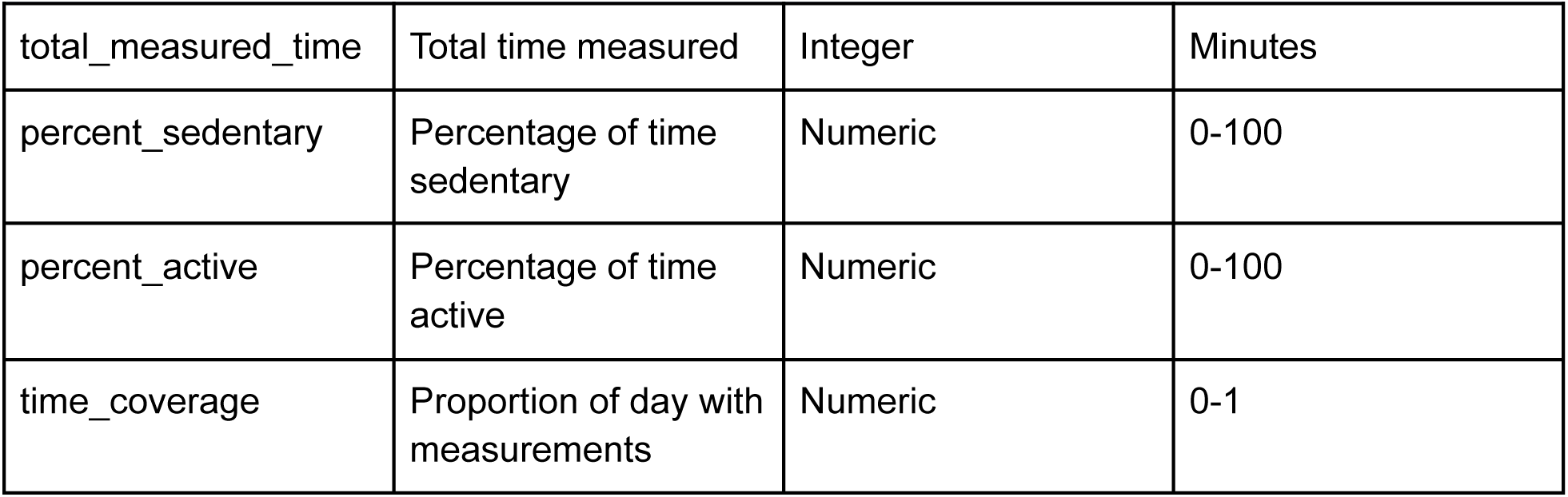
Daily Activity Summary Variables. (N = 24,581 observations)

**Table 7:** PROMIS T-score Variables. (N = 497 observations)

| Variable Name | Description | Type | Values |
| --- | --- | --- | --- |
| STUDY_PRTCPT_ID | Anonymous participant identifier | Character | Format: "P####" |
| Timestamp | Time collecting survey response | Character | "Baseline", "Day30", "Day120" |
| Group | Participant category | Character | "Patients", "Caregivers" |
| t_comp | companionship T-score | Numeric | 0-100 |
| t_glohlth_mh | global mental health T-score | Numeric | 0-100 |
| t_glohlth_ph | global physical health T-score | Numeric | 0-100 |
| t_abroact | ability to participate in social roles and activities T-score | Numeric | 0-100 |
| t_anxty | anxiety T-score | Numeric | 0-100 |
| t_cogfun | cognitive function T-score | Numeric | 0-100 |
| t_deprss | depression T-score | Numeric | 0-100 |
| t_fatig | fatigue T-score | Numeric | 0-100 |
| t_paininf | pain interference T-score | Numeric | 0-100 |
| t_physfun | physical function T-score | Numeric | 0-100 |
| t_slpdist | sleep disturbance T-score | Numeric | 0-100 |
| t_managda | self-efficacy for managing daily activities T-score | Numeric | 0-100 |
| t_managsm | self-efficacy for managing symptoms T-score | Numeric | 0-100 |

**Table 8:** Clinical Outcome Variables. (N = 168 observations)

| Variable Name | Description | Type | Values |
| --- | --- | --- | --- |
| STUDY_PRTCPT_ID | Anonymous participant identifier | Character | Format: "P####" |
| cg_arm | Study arm assignment | Character | 1 = intervention,<br>0 = Control |
| grfs_agvhd_3_4 | grade 3-4 acute GVHD event flag | Character | 1 = occur,<br>0 = not occur |
| grfs_agvhd_date | Days elapsed since transplant until grade 3-4 acute GVHD occurrence | Numeric | 0-100 |
| grfs_cgvhd | systemic therapy-requiring chronic GVHD event flag | Numeric | 0-100 |
| grfs_cgvhd_date | Days elapsed since transplant until systemic therapy-requiring chronic GVHD occurrence | Numeric | 0-100 |
| grfs_relapse | Relapse event flag | Numeric | 0-100 |
| grfs_relapse_date | Days elapsed since transplant until relapse | Numeric | 0-100 |
| grfs_death | Death event flag | Numeric | 0-100 |
| grfs_death_date | Days elapsed since transplant until death | Numeric | 0-100 |
| notes | Additional notes | Character |  |

**Table 9:** Readmission Variables. (N = 198 observations)

| Variable Name | Description | Type | Values/Units |
| --- | --- | --- | --- |
| STUDY_PRTCPT_ID | Anonymous participant identifier | Character | Format: "P####" |
| number_admission | The number of readmission | Integer | 0 = Never,<br>1 = 1st readmission<br>etc. |
| date_admit | Days elapsed since transplant until readmission date | Integer | days |
| admission_reason | The reason of readmission | Character |  |
| date_discharge | Days elapsed since transplant until discharge date | Integer | days |
| days_in_hospital | Number of days in hospital | Integer | days |

**Table 10:** Infection Variables. (N = 221 observations)

| Variable Name | Description | Type | Values/Units |
| --- | --- | --- | --- |
| STUDY_PRTCPT_ID | Anonymous participant identifier | Character | Format: "P####" |
| number_infection | The number of infection | Integer | 0 = Never,<br>1 = 1st infection etc. |
| date_culture_drawn | Days elapsed since transplant until the infected culture was drawn | Integer | days |
| culture_source | The source of culture | Character |  |
| infection_type | The type of infection | Character |  |
| infection_name | The name of infection | Character |  |

## Data Availability

The datasets supporting the results of this article will be made available in Deep Blue Data repository

## Availability of Source Code and Requirements

Project name: dHCT

Github: https://github.com/adityabn6/dHCT

Operating system(s): e.g. Platform independent

Programming language: R

Other requirements: R packages *data.table*

License: GNU GPL

All R scripts for the processing and analysis are available in the subfolder Rscripts

## Declarations

### Consent for Publication

The study was conducted in accordance with the Declaration of Helsinki, as revised in 2013. The study was approved by the University of Michigan Medical School Institutional Review Board (IRBMED HUM#00186436) and registered on ClinicalTrials.gov (NCT04094844). IRBMED-approved informed consent was taken from all the study participants. The U-M IRBMED served as the Single IRB (sIRB) for this multi-site RCT.

### Author Contributions

S.W.C - study conceptualization and finding acquisition, N.E.C, A.H, D.A.H, D.B and T.B - study design and methodology, A.J and R.K - data curation, A.J and N.S - data analysis and visualization, AJ - manuscript writing. All authors discussed the results and commented on the manuscript.

## Acknowledgements

The authors wish to thank the patients and caregivers who participated in this study, along with the study coordinators who made it all possible.

